# Citizen involvement in research on technological innovations for health, care or well-being: a scoping review

**DOI:** 10.1101/2022.11.03.22281892

**Authors:** Catharina M. van Leersum, Christina Jaschinski, Marloes Bults, Johan van der Zwart

## Abstract

Citizen science can be a powerful approach to foster the successful implementation of technological innovations in health, care or well-being. Involving experience experts as co-researchers or co-designers of technological innovations facilitates mutual learning, community building, and empowerment. By utilizing the expert knowledge of the intended users, innovations have a better chance to get adopted and solve complex health-related problems. As citizen science is still a relatively new practice for health and well-being, little is known about effective methods and guidelines for successful collaboration. This scoping review aims to provide insight in 1) the levels of citizen involvement in current research on technological innovations for health, care or well-being, 2) the used participatory methodologies, and 3) lesson’s learned by the researchers.

The search was performed in SCOPUS in January 2021 and included peer-reviewed journal and conference papers published between 2016 and 2020. The final selection (N=83) was limited to empirical studies that had a clear focus on technological innovations for health, care or well-being and involved citizens at the level of collaboration or higher. Our results show a growing interest in citizens science as an inclusive research approach. Citizens are predominantly involved in the design phase of innovations and less in the preparation, data-analyses or reporting phase. Eight records had citizens in the lead in one of the research phases.

Researcher use different terms to describe their methodological approach including participatory design, co-design, community based participatory research, co-creation, public and patient involvement, partcipatory action research, user-centered design and citizen science. Our selection of cases shows that succesfull citizen science projects develope a structural and longtidutinal partnership with their collaborators, use a situated and adaptive research approach, and have researchers that are willing to give-up tradional power dynamics and engage in a mutual learning experience.

## Background

Many technologies are developed to address health, care or well-being related challenges, such as mental health, manage health at home, or coping with an understaffed healthcare sector [1]. These technologies can include new diagnostic or therapeutic methods and devices, ambient assisted living technologies, and eHealth such as monitoring, regulatory, or advisory apps. There is a lot of research into the design, development, implementation, and evaluation of new technologies. However, the scoping review of Krick et al. [2019] showed that implementation of new technologies remains a challenge due to lack of in-depth knowledge about the care environment or users [2].

Citizen science, or the use of scientific principles and methods by non-professional scientists, may be a powerful method to improve public participation in research as well as public health and is regarded as a promising research approach to improve implementation and adoption of technological developments in health, care or well-being by collaborating with the intended users as experience experts [3]. The initial drivers to develop citizen science methods were the increase of research capacity [4], and the possibility to respond more effectively on complex societal problems due to the addition of lay, local or traditional knowledge to scientific knowledge [5, 6]. Besides the positive contributions of citizen science to the scientific processes or knowledge, it has potential advantages for lay people being co-researchers of a project. In health related research as co-researcher could result in a broader understanding of their own health and well-being and acquisition of knowledge about a healthy lifestyle [7]. A list of 10 benefits for co-researchers was developed by Haywood [2013] [8]. These benefits include enhancing science knowledge and literacy, enhancing understanding of scientific methods, improving access to scientific information, increasing scientific thinking, improving interpretation skills, diminishing the gap between science and people, strengthening connections between people and environment, empowering co-researchers, increasing community-building, changing attitudes, influencing policy, and gaining access to broader networks [8]. King et al. [2016] showed similar benefits in several citizen science case studies, where the main benefits concerned scientific literacy [9].

Currently, there is no uniform/ broadly accepted definition of citizen science. So far there is no consensus, and it is even discussed whether a definition is necessary [10, 11]. This non-existence of a definition is accepted and allows stakeholders working in citizen science to use different terms and methodologies [12]. In this paper we define citizen science as research (partly) executed by non-professional scientists, volunteers, citizen scientists or co-researchers. Often the citizen scientists work together or are guided by academic researchers or governmental organisations. Thus, a citizen should be taking active part in any of the research activities and ideally collaborate with professional researchers. We take a similar perspective as Heigl et al. [2019] and exclude projects from our definition that use participants only at as providers and resources of knowledge without any active involvement in scientific activities [11].

Citizen science is an approach which comprises a range of participatory approaches that are embraced as collaborative practices and show promising results [3]. Although citizen science in the healthcare domain is a relatively new and rare phenomenon, it is already an established approach in research fields such as ecology, conservation, and biology [5, 13]. Projects with citizen science approaches attract hundreds of thousands of participants, who are involved in different tasks varying in complexity [14]. The inclusion of citizens is valuable for science [15], for example, when the expertise of the general public or a specific population of citizens is needed to understand and solve a problem [16]. The collective intelligence of the citizens has led to scientific discoveries such as protein folding [17] and air pollution [18]. However, citizens science projects also deal with challenges when it comes to the selection of participating citizens, the needed and available competences of citizens, and the credibility of knowledge gathered by or with the citizens [5, 19].

TOPFIT Citizenlab is a three-year research and innovation program aiming at increasing citizen involvement and participation in researching, testing, modifying and implementing technological innovations for prevention and health promotion. One of the objectives withing Citizenlab is to explore and understand useful methodologies of citizen science in health and well-being research. There is limited knowledge on the benefits of citizen science, and the normative assumptions are often taken for granted [20, 21]. Examples of citizen science do not show a thorough analysis of successes and failures, therefor, little is understood about the advantages or pitfalls [21]. In the field of citizen science, to our knowledge nine literature reviews are performed by Carpini et al. [2004], Irvin and Stansbury [2004], McGuire [2006], Conrad and Hilchey [2011], Garau [2012], Kimura and Kinchy [2016], Kullenberg and Kasperowski [2016], Peter et al. [2019], and Ianiello et al. [2019] [13, 21–28]. These reviews focus on the use of citizen science to encourage public discourse, public decision-making, public management, or environmental research. To our knowledge, there were two literature reviews on citizen science with a focus on health, care or well-being published by Domecq et al. [2014] and Malterud and Elvbakken [2020] [29, 30]. These review focus on patient engagement in healthcare research and show that it is used in many settings. Often the research methods are adapted to make engagement possible for the citizens, and patient engagement is low during the data collection phases or rather tokenistic. Authors of both reviews argue that further research is needed to determine which valuable collaborative methods can achieve more active involvement of patients and define ideas and strategies for citizen science in health research [29, 30].

In our scoping review we will analyse the citizen science methodologies applied in technology development for health, care or well-being. We are specifically interested in which innovation phases citizens were involved, and in the level of citizen involvement. The aim was to provide insight in 1) the levels of citizen involvement in current research on technological innovations for health, care or well-being, 2) the used participatory methodologies, and 3) lesson’s learned by the researchers.

### Research phases and citizen involvement

To answer these questions first, we distinguished three research phases, namely: 1) preparation of research, which included the design of a research, the recruitment process, and design of materials such as interview guides, 2) data collection, which included all (iterative) steps of technology design, data collection and use of feedback throughout these iterations, and 3) data analysis and evaluation, which included methods of analysis as well as reporting of research data and outcome. With this distinction it is possible that one record might describe one, two or three phases in which citizens were involved.

Second, we assessed the level of citizen involvement. We proposed levels of citizen involvement based on the participation ladder of Arnstein [1969] and citizen science terminology defined by Hakley [2013] [31, 32]. Arnstein’s participation ladder is more used in citizen science research to discuss the methodology and possibilities of citizen involvement, for example by van Leersum et al. [2020] about the involvement of citizens in the need of long-term care during the development of a tool for self-assessment, and by Kotus [2013] to explore the level of participation of citizens in the urban policy making of a Polish city [33, 34]. The ladder consists of eight steps including manipulation, therapy, informing, consultation, placation, partnership, delegated power, and citizen control [31]. Each step on the ladder characterizes the role of the participant and the researcher. The higher participants are placed on the ladder, the more influence they will have. This ladder configuration shows that one level is building on the previous level, but there is no logical progression from one to another [35]. The steps manipulation and therapy are considered non-participation. The steps of informing, consultation, and placation are considered tokenism, in which the citizens have superficial involvement. From the informing step in which the citizens are informing the researchers about their personal situation or experiences, towards the placation step in which citizens advise the researchers about needed adjustment, research or innovations. The highest steps of partnership, delegated power, and citizen control are called citizen power, in which the citizens have more and more in-depth control and influence. In partnership the relation and tasks of the citizens and researchers are equal, and with citizen control, the citizens lead the research and researchers are available to support [31].

Hakley [2013] uses four different terms to distinguish involvement in citizen science research [32]. First, crowdsourcing, in which cognitive abilities of citizens are not needed and the citizens use digital devices to collect data that are automatically send and analysed by researchers at a later stage. Second, distributed intelligence, in which the citizens get a certain task such as interpretation or classification of data. Third, participatory science, in which citizens are formulating research questions or problems, and are participating in data collection, but analysis is often done by the researchers. Fourth and last, collaborative science, also referred to extreme citizen science, in which a project is carried out by citizens without researchers being actively involved [32]. Although it might seem from terminology and categorisations of citizen involvement that the higher the level of involvement the better, we will not make this argument in this scoping review. All forms of citizen science could be valuable and often citizen science research could not be defined to one level alone [32, 36]. It should be up to the team *“to determine the best design specifications for their own unique context, enabling citizen science to achieve its full potential”* [10]. Also, as Ferro and Molinari [2009] argue, researchers need to be aware of the complexity of citizens [37]. There is a considerable variation across the population of citizens who might be interested to be involved in different forms. This variation could be in terms of interest as well as educational level or technological skills. For some citizens, these differences make it more difficult or less desirable to participate on higher steps of the ladder. Researchers should be more aware of the variation and should know when to make a step towards the citizens rather than to expect the citizens to move towards them [37].

In our analysis we combine the terminology as used by Arnstein (1969) and Hakley (2013) and define four levels of citizen involvement [31, 32]. The first level is the level of *non-involvement*, this level includes manipulation, therapy, and crowdsourcing. The second level is *sharing knowledge*, including informing, consultation, placation, and distributed intelligence. The third level is *collaboration*, which includes partnership, delegated power, and participatory science, and the fourth level is *citizens in the lead*, including citizen control, and collaborative science. These four levels of citizen involvement will be considered in view of the different research phases (Table 1). Based on the scoping review, we will highlight five cases, and further explore the variation in citizens, how they are involved, and what worked well or what was experienced problematic.

**Table 1.**
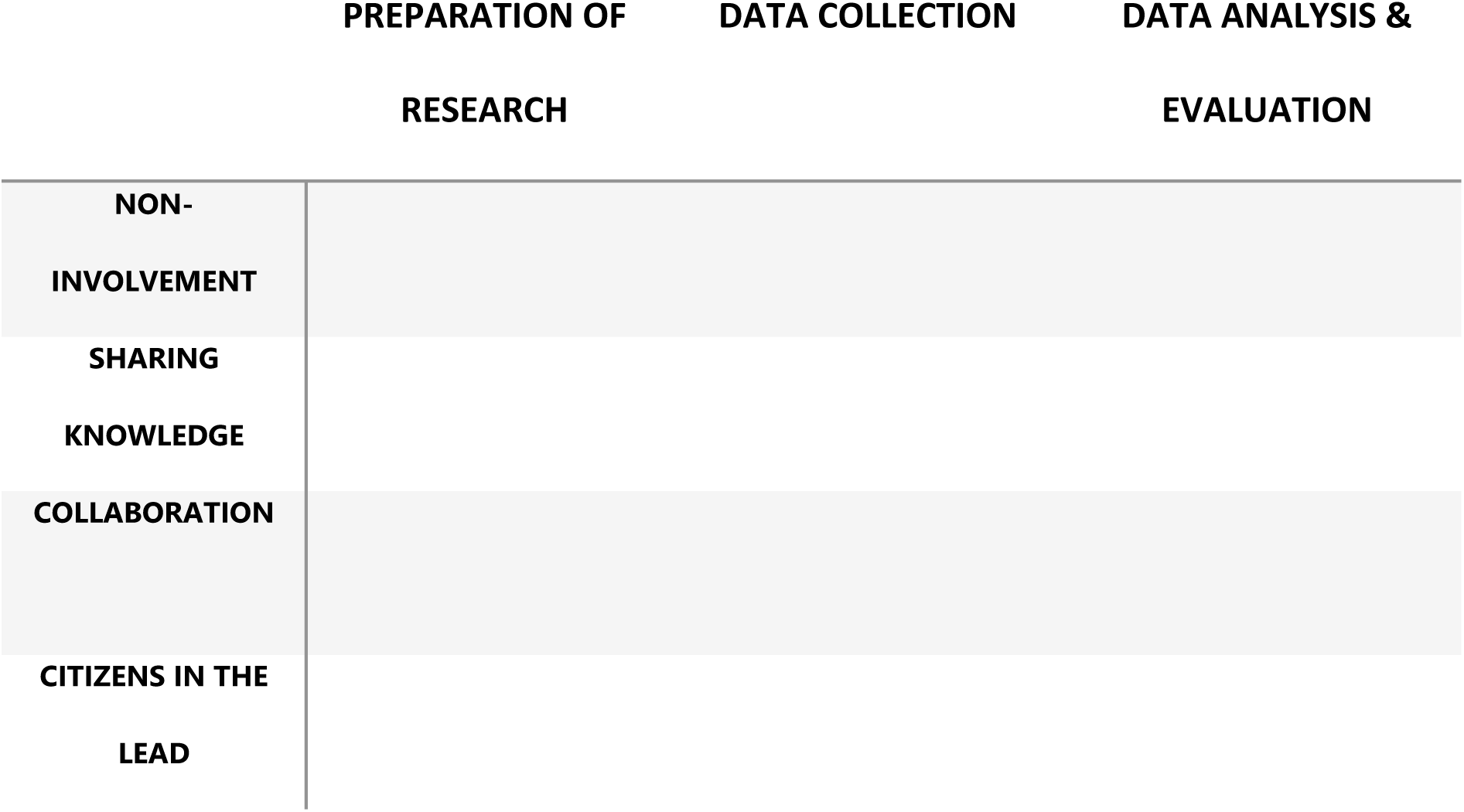
The table used to review the records obtained after the screening of titles and abstracts as part of the scoping review. The levels of citizen involvement (first column) are based on the participation ladder of Arnstein (1969) and citizen science terminology of Hakley (2013) [31, 32]. The phases of research are given in the first row.

## Method

### Methodological approach

A scoping review was performed to examine how citizens are involved in conducted research in the field of health, care or well-being [38]. Scoping review is a reviewing method to map a research area and identify all relevant literature regardless of the study design. This approach is developed to contextualize knowledge and identify the current state of understanding of a specific topic, and to assists in identifying the aspects which are investigated and the aspects which are less or not investigated [39]. A scoping review was chosen due to the broad topic of citizen science and the application of many different study designs within this topic [39]. We investigated citizen science methodologies applied in innovation processes for health, care or well-being. In this article we provide insight in 1) the levels of citizen involvement in current research on technological innovations for health, care or well-being, 2) the used participatory methodologies, and 3) lesson’s learned by the researchers as described in identified records.

### Search strategy

First, we developed a search string that included many combinations of search terms within four research fields: ‘how’, ‘why’, ‘what’, and ‘who’ (Table 2). *How* considers the involvement of citizens, *why* is health, care or well-being related, *what* contains the technological innovations, and *who* are the citizens. For each of these fields specific search terms were defined. For example, the search string for how:

**Table 2.** The main fields of the search string, considering ‘how’ (involvement), ‘why’ (health, care or well-being), ‘what’ (technology innovation), and ‘who’ (citizens).

HOW INVOLVEMENT
|  |  |
| --- | --- |
|  | Citizen science; Participatory research; User involvement; Citizen engagement; Citizen knowledge; Patient and public involvement; Community-based participatory research; Community engagement; Community participation; Citizen participation; Co-creation; Co-design; Participatory design; Co-production; User-centred design |
| WHY | HEALTH AND WELL-BEING |
|  | Health; Healthcare; Care; Wellbeing; Well-being; Lifestyle; Healthy |
| **WHAT** | **TECHNOLOGY INNOVATION** |
|  | Innovation; Technology; App; Digital; Robotics; Robots; Smart; Gaming; Web based; Games; Game design; Tool; Computer-based; Ehealth, E-health; Telehealth; M-health; Mhealth; Mobile health; E-therapy; Digital health |
| **WHO** | **CITIZENS** |
|  | Citizen; Public; Community; Patients; Clients; Elderly; Seniors; Caregivers; Neighbours; Family; Children; Parents; Adults |

> TITLE-ABS-KEY (“citizen science” OR “participatory research” OR “user involvement” OR “citizen engagement” OR “citizen knowledge” OR “patient and public involvement” OR “community-based participatory research” OR “community engagement” OR “community participation” OR “citizen participation” OR “user-centred design” OR “co-creation” OR “co-design” OR “participatory design” OR “co-production”)

As proposed by Jackson and Waters [2005], different combinations of search terms were tested and discussed by the research team and an information specialist [40]. The records acquired with each combination were compared and based on the included and excluded records by addition or removal of search terms, the final search terms were chosen. The final search string was formed by using AND in between the how, why, what, and who, and the terms within each part were connected using OR. To conduct the search, the final search string was entered in January 2021 into the database Scopus.

### Selection of records

For the selection of the records several inclusion criteria and restrictions were chosen. The inclusion criteria were 1) empirical studies, 2) studies describing a method in which citizens were actively involved, 3) health related topics, and 4) technological innovation. Published records were not eligible if:

1. The record did not present the execution of an empirical study. Records describing exclusively the methodology, lessons learned, study design, or reviews are excluded.
2. The used method did not actively involve citizens. Citizens had to be involved to a larger extent than ‘just’ as subjects. Citizens could be professionals if they are the end-users of the technological innovation.
3. The study did not aim to improve health, care or wellbeing.
4. Technological innovation was not part of the record. We used the definition of Behney [1989] *“Medical technologies include the drugs, devices, and medical and surgical procedures used in medical care, and the organizational and supportive systems within which such care is provided”* [41].

Furthermore, we restricted our search to records that were published in the last five years, were peer reviewed or conference papers, and published in English. There were no restrictions on type of research or study design.

The first 200 titles and abstracts were screened by the four researchers. The exclusion criteria for the first screening consisted of no citizen involvement, no health, no technology, and not empirical. The four researchers discussed the differences of inclusion after screening the first 100 records. Then repeated the process for the next 100 records. The remaining records were split and both sets were screened by two researchers, each researcher couple discussed their differences. In case of doubt, the titles and abstracts were also discussed with the other researcher couple.

### Ranking citizen involvement

To classify the used methodologies of the included records, we developed a matrix to rank the citizen involvement within the described research phases (Table 1). The citizen involvement levels were based on the participation ladder of Arnstein [1969] and the citizen science terminology of Hakley [2013] [31, 32]. We defined four levels:

1. Non-involvement: there is no active involvement of citizens.
2. Sharing knowledge: the citizen receives information from the researcher, the citizen provides (advisory) information to the researcher, there is a dialogue in which decisions were made by the researcher.
3. Collaboration: the citizen and researcher are partners, equal in dialogue and equal in taking decisions, the researcher assists in all research activities.
4. Citizens in the lead: the citizen is leading and taking decisions, very minimal assistance of the researcher is provided, only on request of the citizen.

The methods sections of the full text records were screened and ranked on research phase and level of citizen involvement. The first 20 full text rankings were performed by four researchers. A record was directly excluded when it did not fit anywhere in the matrix. The four researchers discussed the differences of inclusion and ranking after reading the first 20 full texts. Then repeated this process for the next 10. The remaining records were divided among the researchers. In case of doubt, another researcher read the full text as well to align the analysis.

### Data analysis

The research phases and the level of citizen involvement was screened. The analysis was based on the descriptions of the methodology in the records. We decided to take only records ranked on the levels of collaboration or citizens in the lead in the final steps of the analysis. The following information was extracted from the records: authors, year of publication, country of research, used methodology, health, care or well-being research topic, technological innovation, number and characteristics of involved citizens, and level of citizen involvement. Besides these details, five records were described in more detail as example cases.

## Results

### Selection and characteristics of records

In total 3846 records were retrieved (Figure 1), no duplicates were found. After screening the titles and abstracts of the records, 2983 records were removed based on the inclusion criteria: empirical study, citizen involvement, focus on health, care or well-being, and technological innovation. Full texts were obtained, and the methods sections of the remaining 861 records were screened and ranked. During methods screening and full text reading 667 records were removed due to no citizen involvement. From the 194 remaining record, 110 records were put-aside based on citizen involvement solely on the levels of sharing knowledge or non-involvement. A total of 83 records were included for further analysis and are presented in table 3. For each record table 3 shows the year of publication, country in which the study was performed, terminology used for the research design, topic of health, care or well-being, technological innovation, involved citizens, and the level of involvement during the different research phases.

**Fig 1.**
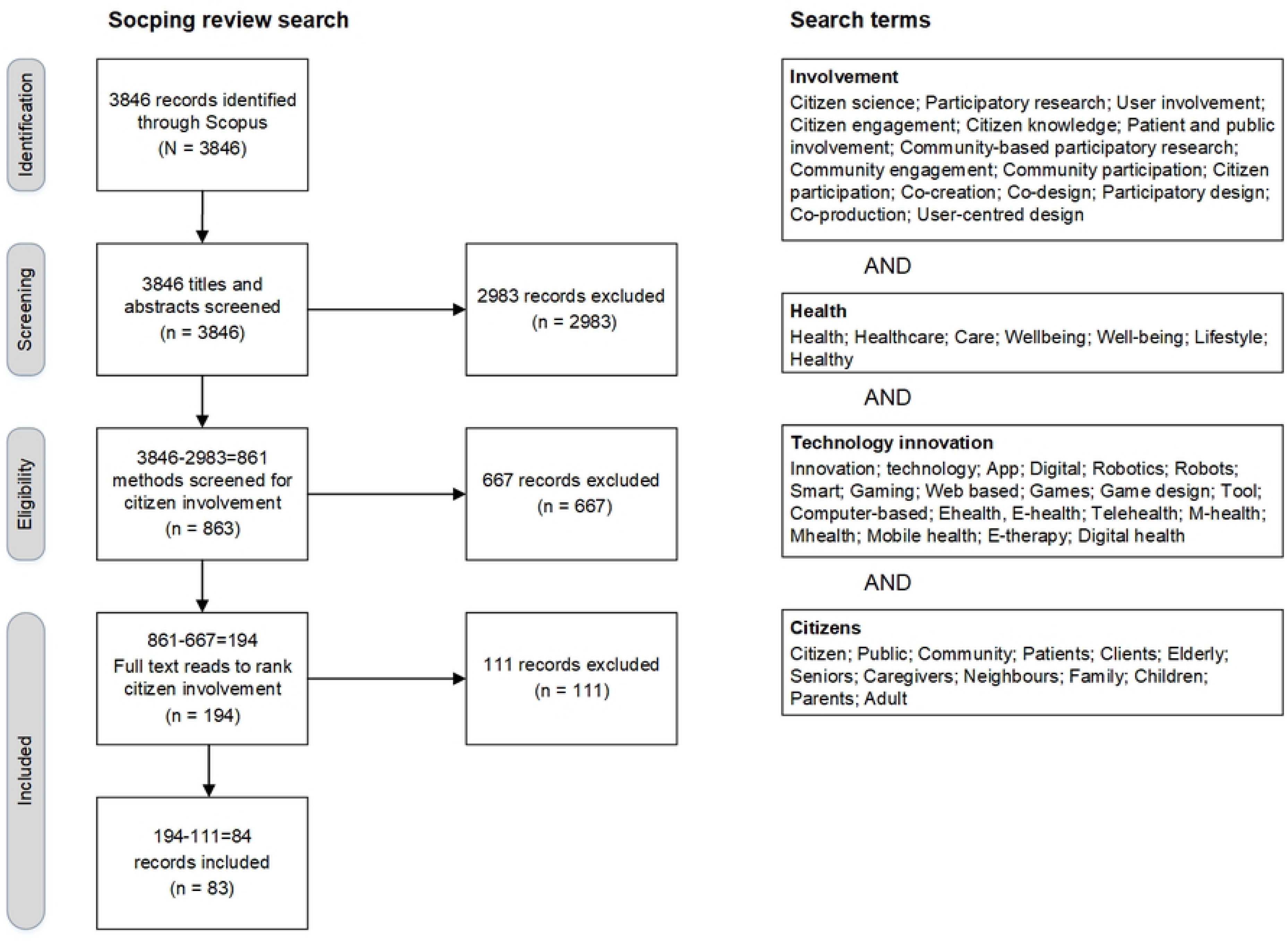
The scoping review search delivered 3846 records with no duplicates. After title and abstract screening 2983 records were excluded. Method screening resulted in the exclusion of 667 records. In 111 records, the study solely involved citizens on the level of sharing knowledge or non-involvement, therefore, 83 records were finally included.

**Table 3.** All 83 included records in alphabetical order. Author, year of publication, country of research, used methodology, health or well-being research topic, technological innovation, number of involved citizens, and the level of citizen involvement in each research phase is distilled from the full texts.

| Authors | Year of publication | Country | Used terminology to describe Method | Health, care or well-being topic | Technological innovation | Involved citizens | Level of involvement in preparation | Level of involvement in data collection | Level of involvement in evaluation |
| --- | --- | --- | --- | --- | --- | --- | --- | --- | --- |
| Abbass-Dick et al. [42] | 2018 | Canada | Participatory design | Breastfeeding | eHealth | 11 Indigenous mothers, 9 care providers | Non-involvement | Collaboration | Non-involvement |
| Abbass-Dick et al. [43] | 2020 | Canada | Co-creation | Breastfeeding | eHealth | 12 young mothers, 8 health care providers | Non-involvement | Collaboration | Non-involvement |
| Ahmed et al. [44] | 2019 | United States | Participatory design | Cardiac resynchronization therapy | Electronic health data | 24 adult patients | Collaboration | Collaboration | Non-involvement |
| Allin et al. [45] | 2018 | Canada | Participatory design | Spinal cord injury | Online self-management tool | 9 persons with SCI; co-designers; key informants | Collaboration | Collaboration | Sharing knowledge |
| Arevian et al. [46] | 2018 | United States | Co-creation | Community resilience | Mobile application | 30 people from the community; co-leadership | Sharing knowledge | Collaboration | Collaboration |
| Arevian et al. [47] | 2020 | United States | Usability study | Obsessive compulsive disorder | Mobile texting apps | 28 patients, 4 care providers | Non-involvement | Collaboration | Sharing knowledge |
| Cesar et al. [48] | 2017 | United States | Community based | Community based health | mHealth technology | 159 people from African | Collaboration | Collaboration | Collaboration |
|  |  |  | participatory research |  |  | American communities |  |  |  |
| De Cocker et al. [49] | 2019 | Belgium | Citizen Science | Public health guidelines | Online survey | 6246 adults | Sharing knowledge | Collaboration | Sharing knowledge |
| DeRenzi et al. [50] | 2017 | India | Participatory design | Community health | Web- and voice-based system | 71 community health workers | Non-involvement | Sharing knowledge | Collaboration |
| Dewa et al. [51] | 2020 | United Kingdom | Co-production | Mental health difficulties | Technology to detect mental health deterioration | 7 young people; co-researchers; advisory group | Collaboration | Collaboration | Collaboration |
| Doty et al. [52] | 2020 | United States | Community based participatory research | Parenting programmes for Latinx families | Mobile app/mHealth | 115 Latinx parents<br>Latinx-serving organisations<br>Latinx community facilitators | Collaboration | Collaboration | Collaboration |
| Edwards et al. [53] | 2020 | United States & Ireland | Community based participatory research | Home-based spirometry in pulmonary fibrosis | Mobile app/mHealth | 36 patients | Collaboration | Non-involvement | Non-involvement |
| Elk et al. [54] | 2020 | United States | Community based participatory research | Palliative care | Telehealth | 14 community members with different ethnicities | Collaboration | Collaboration | Collaboration |
| Erlingsdóttir et al. [55] | 2019 | Sweden | Co-design | Support for home care staff | eHealth services | 16 home care staff from 4 municipalities | Non-involvement | Collaboration | Collaboration |
| Evans et al. [56] | 2016 | United Kingdom | Community based participatory research | HIV testing | mHealth | 12 community members | Collaboration | Collaboration | Collaboration |
| Ferguson et al. [57] | 2018 | United Kingdom | Participatory design | Educational programme for hearing aid users | Interactive video tutorials | 3 hearing aid users and 1 charity advocate people with hearing loss, panel of 11 audiologists | Non-involvement | Collaboration | Sharing knowledge |
| Ferri et al. [58] | 2020 | Italy | Co-creation | Cyber bullying | Massive open online course (MOOC) | 30 children, 2 teachers; co-creators | Non-involvement | Collaboration | Sharing knowledge |
| Fletcher and Mullett [59] | 2016 | Canada | Participatory action research | Healthy lifestyle | Digital stories | 230 younger adults, 14 older adults; development team | Collaboration | Citizens in the lead | Collaboration |
| Franco et al. [60] | 2016 | United States | Co-design | Post-Traumatic Stress Disorder | mHealth/ Mobile app | 32 veterans (peer mentors and mentees) | Collaboration | Collaboration | Collaboration |
| Frauenberger et al. [61] | 2020 | Austria | Participatory design | Social Play for mixed groups of neurodivergent and neurotypical children | Social Play Technologies | 16 neurodivergent and neurotypical children aged 7 to 12, 1 teacher | Collaboration | Citizens in the lead | Sharing knowledge |
| Garcia et al. [62] | 2019 | Brazil | Co-creation | Alcohol and drugs rehabilitation | Digital games | 10 adults with alcohol/drug addiction, 2 healthcare professionals | Sharing knowledge | Citizens in the lead | Collaboration |
| Gardsten [63] | 2017 | Sweden | Participatory design | Self-management of Type 2 diabetes | ICT self-management service | 5 adults with type 2 diabetes, 2 diabetes nurses | Collaboration | Collaboration | Sharing knowledge |
| Garmendia et al. [64] | 2019 | Spain | Co-design | Oncology follow-up | Visualization tool | 2 oncologists and 2 nurses | Non-involvement | Collaboration | Non-involvement |
| Giroux et al. [65] | 2019 | Canada | Co-design | Supporting caregivers of older adults in home care | eHealth | 74 caregivers, community workers, health and social service professionals | Sharing knowledge | Collaboration | Non-involvement |
| Gobl et al. [66] | 2019 | Austria | Participatory design | Social media literacy | Serious game | 71 adolescents | Sharing knowledge | Collaboration | Non-involvement |
| Grant et al. [67] | 2020 | United Kingdom | Co-design, Public and Patient Involvement | Mental health | mHealth | 26 young people | Non-involvement | Collaboration | Collaboration |
| Haufe et al. [68] | 2019 | Netherlands | Participatory action research | Independent living | Gerontechnologies | 19 seniors, 2 technology consultants and 3 municipal community builders | Collaboration | Collaboration | Collaboration |
| Hernández et al. [69] | 2018 | Canada | Participatory design | Force Resistance Training in Hand Grasp and Arm Therapy | Video game | 3 occupational therapists, 1 paediatrician, 12 children | Non-involvement | Collaboration | Non-involvement |
| Hill et al. [70] | 2017 | Canada | Community based participatory research | Stroke education | Educational DVD | 26 indigenous healthcare providers, 108 children | Collaboration | Collaboration | Collaboration |
| Hiratsuka et al. [71] | 2019 | United States | Community-based participatory research | Posttraumatic stress disorder | Website | 6 members of a native community, 5 clinical providers | Collaboration | Collaboration | Collaboration |
| Hong et al. [72] | 2018 | United States | Co-Design | Symptom communication in chronic illness | Digital tracking tool | 13 pairs of chronically ill children and their parents, 11 clinicians | Non-involvement | Collaboration | Non-involvement |
| Jarke et al. [73] | 2019 | Germany | Co-creation | Digital neighbourhood guide | Digital neighbourhood guide | 12 older adults; co-creators | Sharing knowledge | Collaboration | Sharing knowledge |
| Jessen et al. [74] | 2020 | Norway | Participatory design | Personal strengths | mHealth app | 12 patients, 15 care providers, 6 designers, 2 researchers; development team | Collaboration | Collaboration | Collaboration |
| Jessen et al. [75] | 2018 | Norway | Co-design | Self-management of chronic conditions | mHealth self-management app | 22 patients with chronic conditions | Collaboration | Collaboration | Collaboration |
| Jo et al. [76] | 2020 | United States | Co-design | Stress management and mental well-being | mobile self-tracking platform | 5 Korean mothers in the United States | Collaboration | Collaboration | Collaboration |
| Jørgensen et al. [77] | 2018 | Denmark | Participatory design | Rheumatic diseases | e-Device for disease management | 14 patients, 9 care providers | Collaboration | Collaboration | Non-involvement |
| Kanstrup and Bertelsen [78] | 2016 | Denmark | Participatory design | Exercise technology | Exercise technology | 18 young adults with depression or anxiety | Collaboration | Collaboration | Collaboration |
| Kearns et al. [79] | 2020 | Ireland | Public patient involvement | Aphasia rehabilitation | Online user feedback questionnaire | 6 patients with aphasia | Sharing knowledge | Collaboration | Collaboration |
| Kenning and Treadaway [80] | 2018 | United Kingdom | Participatory design | Dementia | Sensory textiles | 13 patients | Sharing knowledge | Collaboration | Sharing knowledge |
| Kildea et al. [81] | 2019 | Canada | Co-design | Patient portal | smartphone app | 445 stakeholders; co-leadership | Collaboration | Collaboration | Collaboration |
| Kim et al. [82] | 2019 | Korea | Participatory design | Autism | Digital self-tracker platform | 5 patients with autism and their partners | Non-involvement | Collaboration | Collaboration |
| LaMonica et al. [83] | 2020 | Australia | Participatory design | Mental health services | Web-based platform | 49 veterans | Non-involvement | Collaboration | Sharing knowledge |
| Lan Hing Ting et al. [84] | 2020 | France | Human-centred participatory design | Fall prevention | Balance assessment tool | 5 elderly people | Collaboration | Collaboration | Collaboration |
| Larsen et al. [85] | 2016 | Denmark | Participatory design | Telemedicine | Shared service centre | 35 clinicians, 29 representatives, 4 patients | Collaboration | Collaboration | Collaboration |
| Latulippe et al. [86] | 2020 | Canada | Co-design | Social health inequalities | eHealth tool for caregivers | 31 community workers, 22 health professionals, 33 caregivers; co-designers | Collaboration | Collaboration | Non-involvement |
| Latulippe et al. [87] | 2020 | Canada | Social justice design | Social health inequalities | eHealth tool for caregivers | 26 community workers, 18 health professionals, 30 caregivers | Sharing knowledge | Collaboration | Sharing knowledge |
| Latulippe et al. [88] | 2019 | Canada | Co-design | Support for older persons | eHealth tool for caregivers | 2 community workers, 2 health professionals, 3 caregivers | Sharing knowledge | Collaboration | Sharing knowledge |
| Lee et al. [89] | 2017 | United States | User-centered design | Support for older adults | Social robots | 5 older adults, 5 mental health therapists; co-designers | Sharing knowledge | Collaboration | Non-involvement |
| Leeming and Thew [90] | 2017 | United Kingdom | Public patient involvement | Patient records | Federated platform | 30 care organisations; steering group | Non-involvement | Non-involvement | Collaboration |
| Leong and Johnston [91] | 2016 | Australia | Participatory design | Aging people | Social robots | 8 older adults | Sharing knowledge | Collaboration | Non-involvement |
| Leslie et al. [92] | 2019 | Canada | User-centred design | Health services | Mobile App to assist caregivers | 25 family caregivers | Collaboration | Collaboration | Sharing knowledge |
| Limaye et al. [93] | 2018 | Peru | Community based participatory research | Maternal mortality | Digital story telling | 135 people from the community | Collaboration | Collaboration | Sharing knowledge |
| Livingood et al. [94] | 2017 | United States | Community based participatory research | Obesity | Digital communication intervention | 30 youths aged 15 to 19 | Collaboration | Citizens in the lead | Collaboration |
| Lo et al. [95] | 2020 | Canada | Symposium | Health informatics | Health information technology | 37 participants | Non-involvement | Collaboration | Non-involvement |
| Lockerbie and Maiden [96] | 2020 | United Kingdom | Co-design | Dementia | Digital quality of life model | 10 care workers | Non-involvement | Collaboration | Sharing knowledge |
| Lu et al. [97] | 2017 | 4 EU countries | Co-creation | Healthy and active ageing | Assistive living technologies | Unknown number of multi-stakeholders | Non-involvement | Collaboration | Non-involvement |
| Maertens et al. [98] | 2017 | United States | Community oriented approach | Human papillomavirus infection | Web-based intervention | 20 Latina young adults, 27 Latino parents; community advisory committee | Collaboration | Collaboration | Sharing knowledge |
| Martin et al. [99] | 2020 | 3 EU countries | Co-design | Healthy lifestyle | mHealth apps | 74 adolescents aged 13 to 16 | Sharing knowledge | Sharing knowledge | Collaboration |
| Matthews et al. [100] | 2016 | United States | Participatory design | Mental illnesses | Smartphone and web app | 3 patient, 3 clinicians | Sharing knowledge | Collaboration | Non-involvement |
| Mburu et al. [101] | 2018 | South Africa | Co-design | Preterm infants | Digital communication | 17 mothers, 17 NICU nurses, 4 doctors | Sharing knowledge | Collaboration | Non-involvement |
| McMillan et al. [102] | 2018 | United Kingdom | Public patient involvement | Health check results | website | 19 members of the public; co-facilitators | Citizens in the lead | Collaboration | Sharing Knowledge |
| McNaney et al. [103] | 2017 | United Kingdom | Co-design | Dementia | Mobile app | 14 young family members of people with dementia | Collaboration | Collaboration | Sharing knowledge |
| Miller et al. [104] | 2016 | United States | Participatory design | Health information | Information technology | 8 parents, 2 children, 3 clinicians | Collaboration | Collaboration | Sharing knowledge |
| Mrklas et al. [105] | 2020 | Canada | Co-design | Knee osteoarthritis | MHealth app | 4 patients, 5 physicians, 3 trainees, 2 decision makers; | Collaboration | Collaboration | Collaboration |
|  |  |  |  |  |  | research coalition |  |  |  |
| Naeemabadi et al. [106] | 2020 | Denmark | Participatory design | Total knee replacement | Telerehabilitation program | 8 patients, 4 health professionals, 4 student assistants, 2 software developers | Collaboration | Collaboration | Non-involvement |
| Payton [107] | 2016 | United States | Co-creation | Health information HIV | Social media | 15 participants | Non-involvement | Citizens in the lead | Non-involvement |
| Rabba et al. [108] | 2020 | Australia | Participatory action research | Autism | Web-based resource | 3 parents, 9 clinicians; advisory group | Collaboration | Collaboration | Non-involvement |
| Rai et al. [109] | 2020 | United Kingdom & Ireland | Public patient involvement | High blood pressure | Digital intervention | 40 patients and partners, 35 care professionals; co-investigators | Collaboration | Collaboration | Collaboration |
| Rathnayeke et al. [110] | 2020 | Australia | Co-design | Dementia | mHealth application | 10 carers | Non-involvement | Collaboration | Non-involvement |
| Reade et al. [111] | 2017 | United Kingdom | Public patient involvement | Joint pain; rheumatoid arthritis | Smartphone app | 20 patients | Collaboration | Collaboration | Collaboration |
| Russ et al. [112] | 2020 | United Kingdom | Participatory action research | Surgical care | Smartphone app | 42 patients; steering group | Collaboration | Non-involvement | Collaboration |
| Sin et al. [113] | 2019 | United Kingdom | Co-design | Psychosis | eHealth intervention | 14 experts; advisory group | Non-involvement | Collaboration | Non-involvement |
| Skovlund et al. [114] | 2020 | Denmark | Public patient involvement | Metastatic melanoma | Dialogue tool | 5 patients | Collaboration | Collaboration | Collaboration |
| Solem et al. [115] | 2020 | Norway | User centred design | Chronic pain | eHealth intervention | 20 project team members, 33 external stakeholders | Collaboration | Collaboration | Collaboration |
| Synnot et al. [116] | 2018 | Australia | Participation in research | Multiple sclerosis | Website | 83 patients; advisory group | Collaboration | Sharing knowledge | Collaboration |
| Tsvyatkov a and Storni [117] | 2019 | Ireland | Participatory design | Type 1 diabetes mellitus | eBook | 24 parents, 20 children; design partners | Non-involvement | Collaboration | Sharing knowledge |
| Tuckett et al. [118] | 2018 | Australia | Citizen science | Physical activity for mental health | Mobile application | 8 older adults aged 65 and older; citizen scientists | Collaboration | Collaboration | Collaboration |
| Vacca [119] | 2019 | United States | Participatory design | Teen-caregiver relationship | Digital media creation platform | 8 Latina teens | Non-involvement | Citizens in the lead | Sharing knowledge |
| Vallentin-Holbech et al. [120] | 2020 | Denmark | Co-creation | Health promotion; alcohol prevention | Virtual Reality (VR) simulation | 9 students; development team | Non-involvement | Collaboration | Non-involvement |
| Van Leersum et al. [34] | 2020 | Netherlands | User-centred design | Long-term care | Web-based preference elicitation tool | 21 end-users; development team | Collaboration | Citizens in the lead | Sharing knowledge |
| Vilarinho et al. [121] | 2017 | 5 EU countries | Co-design | Cystic fibrosis | mHealth application | 14 children, 15 parents, 12 health professionals | Non-involvement | Collaboration | Non-involvement |
| Wikman et al. [122] | 2018 | Sweden | Participatory action research | Cancer | Internet-administered intervention | 6 parents, 2 clinical psychologists; research partners | Non-involvement | Collaboration | Sharing knowledge |
| Yarosh and Schueller [123] | 2017 | United States | Participatory design | Children well-being and resilience | Computing technologies | 12 children; investigators | Non-involvement | Collaboration | Non-involvement |

From the 83 included records, eight were published in 2016, 14 in 2017, 15 in 2018, 17 in 2019, and 29 in 2020. Most studies were conducted in the United States (N=17) [44, 46–48, 52, 54, 71, 72, 76, 89, 94, 98, 100, 104, 107, 119, 123], followed by Canada (N=14) [42, 43, 45, 59, 65, 69, 70, 81, 86–88, 92, 95, 105], the United Kingdom (N=12) [51, 56, 57, 67, 80, 90, 96, 102, 103, 111–113], Denmark (N=6) [77, 78, 85, 106, 114, 120], and Australia (N=6) [83, 91, 108, 110, 116, 118]. Five studies were conducted in multiple countries [53, 97, 99, 109, 121], three studies were conducted in Sweden [Erlingdottir et al. 2019; Gardsten 2017; Wikman et al. 2018], and Norway [74, 75, 115], and two in Austria [61, 66], the Netherlands [34, 68], and Ireland [79, 117]. One study was conducted in Belgium [49], India [50], Italy [58], Brazil [62], Spain [64], Germany [73], Korea [82], France [84], Peru [93], and South Africa [101].

One of the 83 records was based on a quantitative research design, all other included record had a research design based on qualitative methodologies. Participatory design was most prominent (N=24) [42, 44, 45, 50, 57, 61, 63, 66, 69, 74, 75, 77, 78, 80, 82, 83, 85, 91, 100, 104, 106, 117, 119, 123], followed by co-design (N=16) [55, 64, 65, 72, 76, 81, 86, 88, 96, 99, 101, 103, 105, 110, 113, 121]. Other often used terms were community based participatory research (N=9) [48, 52–54, 56, 70, 71, 93, 94], co-creation (N=8) [43, 46, 58, 62, 73, 97, 107, 120], public and patient involvement (N=7) [67, 79, 90, 102, 109, 111, 114], participatory action research (N=5) [59, 68, 108, 112, 122], and user centered design (N=4) [34, 89, 92, 115]. Citizen science was only used in two records [49, 118], and usability study [47], co-production [51], human centered participatory design [84], social Justice design [87], symposium [95], community oriented approach [98], and participation in research [116] were all only used once to describe the research design.

Besides the term citizen, participant, patient, actor or (end) user, there are different terms used to describe the involved citizens. These include key informants [45], co-leadership [46, 81], co-designers [45, 65, 86, 89], co-researchers [51], co-creators [58, 73], co-facilitators [102], co-investigators [109], inventors [123], design/research partners [117, 122], citizen scientist [118], community advisory committee [98], research coalition [105], steering group [71, 90, 112], (expert) advisory group [51, 108, 113, 116], and development group/team [34, 59, 74, 120]. Fifty-nine of 83 records included adults, 11 records included younger adults (13 to 25 years old) [43, 51, 58, 59, 67, 78, 94, 98, 99, 119, 120], eight records included children (12 years or younger) [61, 69, 70, 72, 104, 117, 121, 123], and nine records included older adults [59, 68, 73, 83, 84, 89, 91, 97, 118]. The number of involved citizens is variable among all records, most studies involved between 30 and 40 citizens, with a highest amount of 6246 [49] and the lowest of five citizens [76, 82, 84, 114].

### Research phases and citizen involvement

Full texts were read of 194 records. A total of 111 records involved citizens solely on the levels of sharing knowledge or non-involvement. In these records researchers were in the lead during all research phases and decisions. Sixty-five records were considered to have citizen involvement on a collaboration level in one or more research phases. There were eight records analysed as having citizens in the lead in one research phases. These records had citizens who were responsible and made decisions regarding a development process or were themselves the initiators of a project. Most records involved citizens during the data collection phase. Forty of 83 records had user inclusion in all three research phases, 11 records involved citizens in the data collection and evaluation phase, ten records in the preparation and data collection phase, and one record in the preparation and evaluation phase. Thirteen records described citizen involvement only in one research phase. Table 4 shows the level of citizen involvement in the different research phases of all 194 included for full text reading. Five records are further discussed in the next section as example cases of citizen involvement in research regarding health, care or well-being.

**Table 4.**
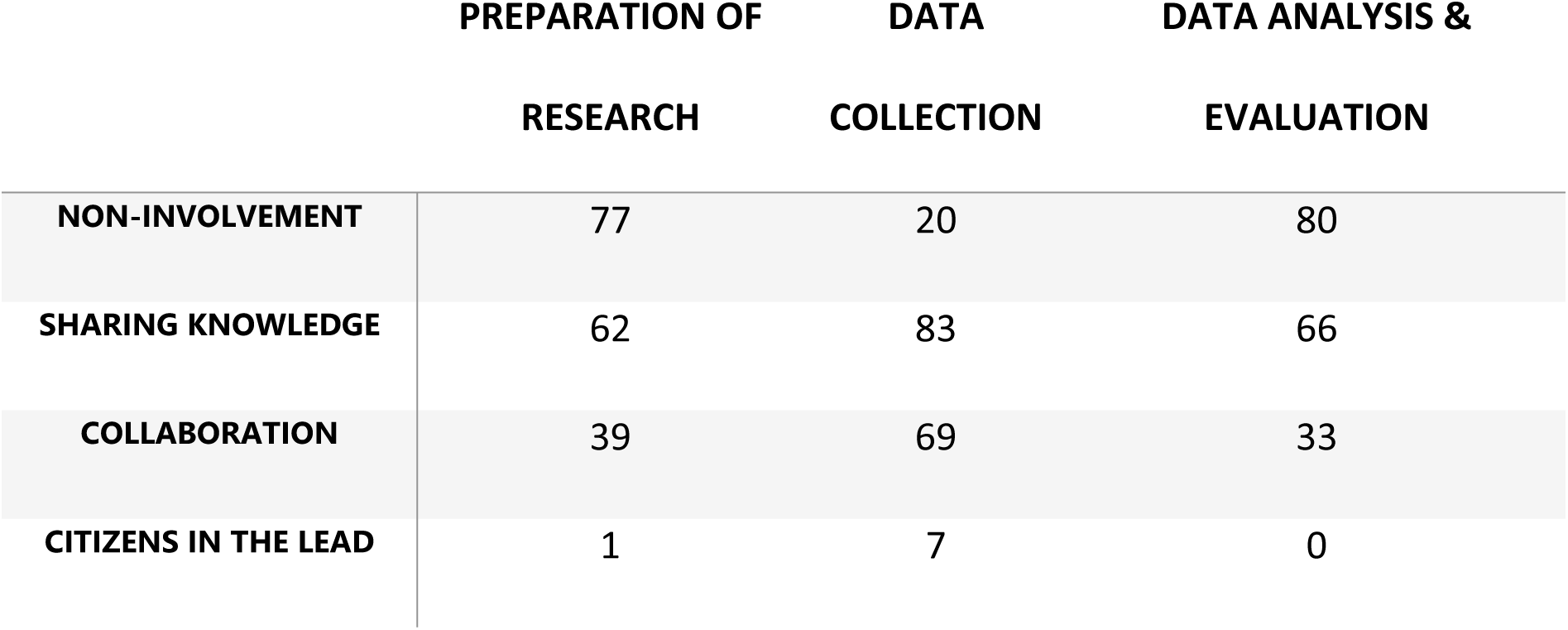
Number of records for each level of citizen involvement and within each research phase. This table is based on the 194 records included for full text reading.

|  | PREPARATION OF<br>RESEARCH | DATA<br>COLLECTION | DATA ANALYSIS &<br>EVALUATION |
| --- | --- | --- | --- |
| NON-INVOLVEMENT | 77 | 20 | 80 |
| SHARING KNOWLEDGE | 62 | 83 | 66 |
| COLLABORATION | 39 | 69 | 33 |
| CITIZENS IN THE LEAD | 1 | 7 | 0 |

### Example cases with high level of involvement in multiple phases

I In this section five example cases are elaborated upon to show a diversity of topics, methods, and citizen inclusion. These cases are chosen based on inclusion of citizens in multiple research phases and at a level of collaboration or higher in one or more research phases. The aim of these cases is to analyse the citizen science method during the innovation process in the domain of health, care or well-being. How were citizens involved, who were involved, what worked well, what kind of problems were met, and what kind of motivation do researchers or citizens have?

#### Case 1. An online self-management tool for spinal cord injury [45]

The technology in this case is an online tool to promote self-management in order to avoid rehospitalization of patients after spinal cord injury (SCI). The data was collected in 2015 and 2016 involving Canadian citizens of different provinces. This case is chosen because it shows a clear description of the organisation and collaboration between the different stakeholders. The authors included several teams demonstrating a divers and broad network (see Box 1 for a case description).

##### Box 1. Case description: Online self-management tool for spinal cord injury

Self-management tools are available for many chronic conditions. The use of these tools is linked to a decrease in rehospitalization rates. However, satisfaction with most self-management tools is low, and there is no known online tool for users with SCI. The goal in this study was to apply a participatory design process in order to develop an online self-management tool for users with SCI. This process was chosen by the authors to define design constraints and solutions. Sixteen individuals, seven researchers and nine persons with SCI, participated in ten different meetings of the participatory design process. Experiences with self-management were discussed in the first meetings (exploration). Thereafter, features for a new online tool were defined (discovery). These features were translated and embedded in a prototype which the group discussed in following meetings, and feedback was used to make iterations (prototyping). The design process and collaboration with potential users was valuable to engage end users in the design of a tool to promote self-management after SCI.

In this case, the participatory design process consisted of three steps: exploration, discovery, and exploration [124]. The design process was described as an interactive design in which the knowledge of end-users was acknowledged and taken into the design by involving users. The process should include expressing tacit knowledge and encouraging sensitivity [45]. The design process included a core co-design and co-development team meeting on regular basis. The record clearly shows the connections between this core team and the council. This council consisted of a consumer advisory group of users with SCI and a product advisory group of researchers and a clinician [45]. Several other records described a collaboration between different groups, for example Wikman et al. [2018] included parent research partner group and an expert research partner group [122], Russ et al. [2020] made use of a steering group in which public representatives collaborated with researchers [112], and Mrklas et al. [2020] had a research coalition [105]. A difference with the study of Allin et al. [2018], quite often these advisory boards did not include the actual users, but healthcare professionals [45]. The minimal involvement of citizens in the evaluation phase could be seen as a limitation of this case, but in all other phases it was very strong. Furthermore, they recognize possible bias due to a low number of involved citizens, and we noticed a lack of involvement of citizens with a lower educational level. Allin et al. [2018] discuss that all involved citizens had access to high-speed internet, which is associated with a high self-reported health status, and therefore these involved citizens are maybe less in need of self-management than those who are not online [45].

Beforehand the authors motivated their choice to use a participatory design, because it might empower and educate the citizens, and encourages them to adopt the developed application. The three benefits reported afterwards include *“elicitation and consideration of diverse accessibility considerations”*, *“prioritization of features and identification of core design concerns”*, and *“co-creation of acceptable strategies and techniques to mitigate identified concerns”* [45]. Although the expected benefits considered the citizens, the experienced benefits show the value for the development process.

#### Case 2. Digital stories for health promotion [59]

In this case, digital stories are the technologies, which are at the same time the technologies under development as well as the technologies used to perform the research. Sharing knowledge on healthy lifestyle and increasing self-esteem are the health goals with the stories. From 2012 to 2014, 18 workshops on digital stories were organised in communities from Victoria to Ahousat in Canada. This second case was chosen due to intensive groupwork and collaboration between younger and older adults within the community as researchers. A mutual learning process was present between the younger and older adults to develop the digital stories (see Box 2 for a case description).

##### Box 2. Case description: Digital stories for health promotion

Aboriginal youth in Canada experience lower health and well-being due to different challenges they face. Digital stories can emphasize on community connection and cultural continuity, which are factors influential to improve health and well-being. The aim of the study was to create and share digital stories to support health promotion and encourage interactions within a community. This creates a place for younger and older adults where knowledge on healthy lifestyle could be shared, where mentorship could be facilitated, and where self-esteem could be increased by celebrating identity and community. Participatory action research was used to engage younger and older adults in the community. A youth research team was formed with eight younger adults. Next to this team there were 60 core younger participants, 170 younger workshop participants, and 14 older adults involved. At the start, the youth research team was trained to create digital stories. After workshops in which the older adults taught about their healthy lifestyle, the younger and older adults together created mini stories with the use of cameras, audio recorders, and story boards. These stories were evaluated during follow-up sessions, and a small group of younger adults were trained to lead other communities in creating their digital stories.

Fletcher and Mullett [2016] refer to McIntyre [2007], but do not give a definition for their research method [59, 125]. The authors argue that participatory action research was beneficial because it was a non-prescriptive approach. Other records using similar participatory action research methods do give a definition. For example Wikman et al. [2018] based their design on Kindon et al. [2008], “*which is a collaborative process of knowledge production and co-learning, placing people with lived experience at the centre of the process*” [122, 126], and Rabba et al. [2020] based their method on Baum et al.[2006], “*the stakeholders are integral to all parts of the research process, participants’ active role in the outcome sets participatory action research apart from traditional research methods where the participants tend to be more passive in their receipt of research outcomes*” [108, 127]. Often a participatory action research is divided in phases. Rabba et al. [2020] used planning, action, observation, reflection, and using the new learning to plan further steps in their research [108], and Haufe et al. [2019] used the two phases of understanding the current and developing a tool [68]. All participants of the study by Fletcher and Mullet [2016] were involved in the phases planning, designing, and evaluation of the community workshops [59].

The research process gave the community members together with the researchers the opportunity to create a unique, broad, and holistic concept of healthy lifestyle, including personal wellness in body, mind, and spirit. Clear outcome and follow-up include ownership of the stories by the communities, the youth took on leadership in training other communities to develop digital stories, and the older adults took a teaching role in ecology, traditional food preparation, cleansing rituals, and medicinal plants [59]. There were more records discussing the need for public health interventions and the involvement of community members in creating digital stories based on their community and personal stories. Although in most records the creation was done by the community members, this creation was more seen as a needs assessment activity and the researchers chose which stories were used in discussions [93].

It seems that the aim to use a participatory action approach with digital stories to improve health and well-being was positive. The creation of stories by community members was relevant to capture a personal voice and made explicit which public health themes were recognized. The workshops were fun and engaging, and the youth experienced a process with the older adults giving them a voice and a sense of belonging. However, Fletcher and Mullett [2016] also acknowledge that the participants were very active in the creation of stories, presenting, training, and workshops, but there was a much lower interest in planning and analysis [59].

#### Case 3. Design social play things [61]

This case is about technology for social play for mixed groups of neurodivergent and neurotypical children. The well-being goal is to scaffold and support the development of complex social skills through technology-facilitated social play. In the ‘SocialPlayTechnologies’ project the researchers collaborated over the course of three years with two inclusive mainstream primary schools in Vienna, Austria. Three social play technology prototypes were designed together with three groups of children, aged 7 to 12 years. We selected this case due to researchers’ effort to give children the lead in the collaborative design process, their flexibility in adapting to the children’s needs, and to showcase how citizen science can be executed with challenging target groups (see Box 3 for a case description).

##### Box 3. Case description: Design social play things

Social play is an essential part of children’s healthy development. For neurodivergent children engaging in social play with neurotypical peers that have different interaction styles can be challenging, often leading to a preference for solitary play. This study sought to explore how digital technologies can facilitate and support social play among mixed groups of neurotypical and neurodiverse children. The aim was to create technologies that are engaging for both target groups, interactive, open-ended, robust, embedded in their natural play context, extendable, and non-normative. The researchers engaged in a long-term participatory design (PD) process with three groups of children aged 7 to 12 years in. In total 16 children participated as co-designers in this study. The PD process consisted of weekly or bi-weekly series of design workshops (50 in total across the three groups) with at least two research team members as facilitators. Groups were completely free in what social play technology they would design. The first phase of the PD process focussed on exploring children’s perception of playfulness. This phase was followed by various technology immersion activities to introduce the children to sensors and actuators. In the next phase, researchers provided new input through material, scenarios, and narratives, while also narrowing the design space to agreed-upon ideas. When concepts were more concrete, researchers started to bring in first prototypes or groups collaboratively designed prototypes. Three distinct social play technologies were designed: 1) a concept with coloured fabrics embedded with interactive lighting called LightSpaces, 2) a set of interactive pads that trigger sounds called MusicPads, and 3) a concept consisting of a reading lamp with a Raspberry Pi, a camera and a projector called PictureStage. Towards the end of the design process researchers worked on increasing the maturity of the prototypes for the purpose of evaluation. This evaluation phase is escribed in another paper.

The researchers applied a PD to reach their design goal of creating social play technologies suitable for the different ways neurodivergent and neurotypical children make sense of the world, can only be achieved by including these children directly in the design process [61]. As neurotypical adult researchers they felt unequipped to imagine the children’s distinct sense making process in social play. Similar reasoning on the value of PD is given by [42] who argue that PD ensures that a design solution meets the target groups needs and beliefs, and is culturally relevant. Besides understanding perspective and producing relevant design solutions, PD can provide other benefits to participants, such as fostering agency and teaching useful skills [123]. The researchers’ design process was built on previous PD work with children including co-operative Inquiry [128]. Co-operative inquiry has three key elements: 1) children are regarded as research partners and involved throughout the design process, 2) design activities with low-tech materials, and 3) technology immersion in which children explore the design space of novel technologies. Yarosh and Schueller [2017] also worked with children and showed methodological similarities by having co-operative inquiry as a framework to build their PD process, and having long-term engagement since they met with the children twice a week for 14 design sessions [123].

Another concept, central in the researchers’ design process, is ‘Handlungsspielraum’ (in English: room to act) [129]. To open up Handlungsspielraum it was important for the researchers to create design activities that balance structure and freedom through the used materials, planned activities, physical setting and role of the facilitator. While structure provides safety, guides participants, and offers stimuli to participant, freedom allows participants to explore their creativity and ideas. Researchers found that this balance between structure and freedom had a similar effect on creativity within play. Hence, this balance was also reflected in the final prototypes through modularity, manipulability and the option to use the prototypes without the technological component. The researchers in this case tried to let children lead the design process. This is reflected in the fact that each group came up with a different social play technology. Moreover, while the design process followed the same overall blueprint across design groups, the overall pace and specific content of each design session was different for each group and evolved based on the children’s interest and conceptions of playfulness. On the other hand, the researchers noted that they sometimes made design decision contrary to the wishes of the children to limit the complexity of the designed technologies.

Researchers in the current case indicated that working with a mixed group of neurotypical and neurodivergent children was challenging, especially in the groups where no teacher was present. Researchers had to act as mediators when conflict occurred between children and prepared for this task by drawing on previous literature about appropriate design frameworks for this target group. Moreover, they interviewed the teachers at the start of the design process to learn about the group dynamics. Some team members received a training from a special needs educator to learn how to deal with conflict.

#### Case 4. New voices to design exercise technology [78]

The technology in this case is a health promoting digital application to support vulnerable young adults in exercising and physical activities. The ‘Pulse Up project’ is a collaboration of a general practitioner (GP), community workers, and citizens in a neighbourhood in Denmark. Eighteen young adults, aged 18-30, suffering from depression and/or anxiety, and having a low level of physical activity, participated in the exploration and manifestation of design ideas. This case is chosen because of the situated PD process, the creative PD activities, and the inclusion of vulnerable citizens in the choice of research activities, design ideas, analysis, and evaluation of each step in the process (see Box 4 for a case description).

##### Box 4. case description New voices to design exercise technology

Most research into digital technologies to support exercise is centred on people already engaged in exercise activities, while those who are challenged to exercise and are at risk of ill health due to low physical activity level have a limited voice in technology design. This research presents a PD process with young adults suffering from depression and/or anxiety, living in a neighbourhood identified as a high-risk health zone, and motivated but faced with challenges to exercise. The design process consisted of two interventions. The first intervention focussed on developing appropriate participatory methods for bringing new voices to the design of exercise technology and to gain initial insight to inform future design. The second intervention focussed on exploring and manifesting design ideas. Participants met twice a week for eight weeks in the neighbourhood’s local gym to exercise with an instructor and participate in a total of 16 PD activities. Eleven young adults participated in the first intervention. Four continued in the second intervention, and seven new participants were recruited. Exercise itself was not challenging, but the challenge was in the sub-activities to complete before exercise. The participants created three visions to meet with this challenge: 1) dragging friends to exercise, 2) keeping an eye on the activities of group members, and 3) a ‘join’ functionality as an easy way to join activities. The participants’ visions on exercise technology were low since they did not prioritize tracking and competition, health information or exercise instructions. However, based on their challenge, they created visions for digital support.

To bring the voices of vulnerable young adults into the design process of digital technologies to support health promotion, Kanstrup et al [2016] chose for a situated PD [78]. The process combined physical exercises with PD activities. Although the researchers initially planned to do the PD activities in a classroom setting, the participants expressed a dislike of classrooms and preferred activities in the gym. Therefore, the PD activities were integrated with the physical activities in the gym. To change the setting, they made rules for the activities to emphasise on the need for professionalism, effectiveness, and the ability to trigger design. This formed the foundation for the development of a design cycle with progressive stages, supported by game-like PD activities, continuous analysis and evaluation of results. All activities were developed as a fusion of training sessions with music and exercises, and PD activities. In addition, Kanstrup et al. [2016] developed a variation of artefacts to trigger exercise and technology design [78].

The previous case had a similar approach in their research of social playthings. Both the location and the integration of playful activities became part of the design process. Yarosh and Schueller [2017] also reported that they deviated from their original plan of putting the researchers in charge of video-recording design sessions [123]. They decided to give the participants control over the recording process to change the perceived power-imbalance between researchers (as data-collector) and participants (as data-objects). These studies show the benefits of situated research in which group activities are part of the research process. Although this situated approach can be in a physical place, it can also be interpreted as a safe space for open discussions, as is visible in the study of Dewa et al. [2020]. In their study with young people with mental health, a WhatsApp-group became the place for open discussions between the co-researchers [51].

Each PD intervention was evaluated by the participants, who were engaged in an iterative analytical process. Visualisations of analytical results were shared with the participants between two interventions, and they were invited to reflect on the presented analysis and to reconstruct any misinterpretations, missed insight or future visions. After the second intervention, the co-researchers conducted a final analysis of the visions, in which they identified design elements for exercise technology. The created visions move beyond an understanding of exercise technology for vulnerable groups as simply information apps, towards an ambition to design mediators of community health resources in which digital communication is the mediator for physical activities. This requires critical insight about adolescent use of digital communication and the potential importance of messaging, apps, gaming, wearable technology, and rapid changes in youth communication and use of digital technology in developing adolescent physical activity health promotion. This is visible in the study of Livingood et al [2017] in which they develop a digital communication intervention to reduce adolescent obesity [94].

#### Case 5. A person-centred patient portal [81]

This study focuses on a patient portal app for patients with cancer. A patient portal is a secure extension of an electronic medical record (EMR) of a health care institution where patients and professionals have access to [130]. In the study of Kildea et al. [2019] participatory stakeholder co-design was applied, involving patients and health care providers [81]. Data were collected between 2015 and 2018. This study was selected because of the interface between patients and the medical world, and the authors strongly show their used participatory stakeholder co-design (see Box 5 for a case description).

##### Box 5. Case description Design and development of a patient portal (eHealth)

Patient portals represent a real-world example of patients facing electronic health (eHealth). Patient portals become important in daily care because they provide patients with personal health information and contribute to patient engagement and empowerment. A participatory stakeholder co-design was applied involving patients and health care providers. Six core elements were used: equal co-leadership, patient preference determination, security, governance, and legal input, user evaluation and feedback, continuous staff input, and end-user testing. Regarding ‘equal co-leadership’, all three co-leads, a patient, a radiation oncologist, and a medical physicist, were equally involved in decisions and in constant communication. ‘Patient preference determination’ was conducted by a voluntary convenience sampling survey, to obtain input from people receiving cancer treatment. For ‘security, governance and legal input’ purposes a Security and Governance team provided guidance regarding the security and confidentiality of patient data and compliance with applicable regulations. Three patients were participated in a focus group to provide ‘user evaluation and feedback’. The prototype was demonstrated, and participants were observed. A second focus group was conducted before pilot release, consisting of five members of the cancer centre’s patients committee. ‘Continuous staff’ input was ensured by a clinical co-lead, radiation therapists, staff ranging from health care providers to senior management, medical physicists, oncology nurses, radiation oncologists, administrative assistants, and the board of directors of the institution. Finally, to ensure ‘end-user testing’, students and real patients tested the first version of the prototype.

In this case the differences between co-design, patient-centred design and person-centred design were described. In co-design, patients help identify the process or project that needs to be designed based on personal experience. Patient-centered design focuses on fulfilling the needs of the patients, but the project may not have been identified by patients or involve patients. Person-centered design focuses on patients’ need as a whole person and as an equal partner in their care [131, 132]. Kildea et al. [2019] described that the term patient co-design is often confused with patient-centered design and person-centered design [81]. Therefore, the term participatory stakeholder co-design was preferred as term in this study. Patients and clinicians were involved as equal partners, and both actively participate in all parts of the design process [81]. Some other studies involving both patients and health care providers used the term PD [42, 57, 74, 77, 85, 100, 104, 106, 108].

Furthermore, in this case an agile development approach was applied, which consisted of rapid prototyping and testing of various features [133]. A team of developers, students in computer science and medical physics, and co-leads worked together. This led towards a person-centered, clinician acceptable, and informatics feasible patient portal. Comparable to this case, in the study of Leeming and Thew [2017] a patient data platform was developed involving different stakeholders as part of a steering group. Public and patient views were explored during design sessions, and the steering group highlighted the key learnings from the feedback to guide the design [90].

Kildea et al. [2019] described several challenges during the process of this study [81]. For example, the equal co-leadership, where patients are on an equal level as clinicians. This was often not clear to people outside of the immediate design and development team because the hierarchical health care system assumed that projects must be led by a clinician. Also, challenges regarding security, governance, and legal input, especially time-consuming legal issues, were encountered, for example intellectual property, liability, and the contents of the patient disclaimer form. However, as a result of this study a final design of the patient portal smartphone app for patients with cancer was developed from scratch within the health care system based on the principles of person-centeredness design.

## Discussion

### Main findings

The aim was to provide insight in 1) the levels of citizen involvement in current research on technological innovations for health, care or well-being, 2) the used participatory methodologies, and 3) lesson’s learned by the researchers as described in identified records. From the 194 records included after full text reading, 83 records involved citizens on a collaboration level in at least one research phase. The level of citizen involvement differed across research phases. In contrast to the review of Domecq et al. [2014] in which citizen involvement was lacking or tokenism in the data collection phase, in our scoping review most citizens were involved in the data collection, and less in preparation or evaluation [29].

The metrics of these 84 records show that there was a two-fold increase in one year between 2019 and 2020. This shows a growing interest in citizen science for health, care or well-being in the last years. Health, care or well-being research was originally considered less adequate for citizen science, but exceptions are arising [13]. The increase is also visible in the scientific journals that are picking up citizen science as a separate field. The journal ‘Citizen Science Theory and Practice’ was a first journal on citizen science, with a first issue in 2016 [134]. Other journals, such as PlosONE, include citizen science as a search or subject area. In the domain of health, care or well-being the use of citizen science was also incorporated recently, considering the entry of the Medical Subject Heading (MeSH) in 2020. Looking at the countries where the citizen science studies were performed, most of the 83 records in our sample originated from developed countries. In the entire dataset (N=3846), there is a lot of community-based research in developing countries, and in most research the citizens are involved, but most records were excluded due to lack of technological innovation [135–138]. The included studies show a variation in the number of citizens involved, but most studies did not include more than 20. Although the number of citizens in the studies is often argued as sufficient or data saturation was reached, it introduces the risk of recruiting only a minority of the population, often the well-educated and most affluent citizens [37].

Apparent from all records was the terminology used to describe the research methodology or the citizen involvement. The term citizen science was only used in two records and participatory design was most prominent in the included records. Identical authors give different terms to their research design across studies [42, 43, 46, 47, 86, 87], and when using similar terms, the authors base the research method on different sources [86, 88]. Inclusion of all the different research methods is due to our broad search string, however, this is in line with continuing discussions on the definition for citizen science [11], and the acceptance of working in citizen science with different terms and methodologies [12]. Den Broeder et al. [2018] also wrote *“some approaches in health research strongly resemble citizen science, one of these is participatory action research”* [5]. However, they argue that there is a main difference between participatory action research and citizen science, and that difference is in the action part, which refers to the need to act, having change, addressing a specific problem, or developing an intervention. Citizen science can also be a method used to do research without a focus on any action. Furthermore, participatory action research aims towards a strong involvement of citizens and in citizen science, the citizens can be engaged in a less intensive way [5]. Regarding the two records naming their research method citizen science, the level of involvement was diverse. One aimed to reach a large audience with a citizen science-mass experiment with no focus on action [49], and the other trained citizen scientists to become agents of change in their own environment which had a focus on action [118]. These examples show that there is no consensus on the use and terminology of citizen science and related methodologies.

These different views and approaches towards citizen science raise questions regarding the value and benefits of citizen science. Greenwood et al. [2019] argue that active involvement of citizens in the preparation phase ensures that the most relevant research questions will be asked [139]. Also, successful involvement of citizens causes sensitivity and understanding of needs to incorporate in design and implementation [61, 140]. Thereby increasing the chance of adoption and sustainable implementation. Many records in this scoping review consider a form of citizen science to raise awareness on health, care or well-being issues. This is in line with the benefits of citizen science for the citizen scientists [8, 9]. However, little evaluation was done in the included empirical studies. Some discussed the possible benefits for the citizen scientists, such as improved personal health. However, these issues were not obtained from the citizens, but expected or experienced by the researchers. One record discussed that the development team had the feeling of being heard and all ideas were considered in the development [34], but it is often unknown how citizens have experienced their contributions and if they would recognize their ideas as part of a prototype [141].

Compared to traditional science, citizen science asks for different research approaches and skills from researchers. The selected cases imply that successful citizen science project develope a structural and longtidutinal partnership with their collaborators [45, 59, 61, 78, 81]. Citizen science requires an open attitude, flexibility and context sensitivity from researchers. They have to give-up traditional power dynamics, step away from their own expectations and pre-conceptions, be sensitive to the signals of their co-researcher and be willing to engage in a mutual learning experience. For example, researchers might have preconceived theoretical knowledge about the topic of interest and probably about the research population, but this does not always align with the views of the co-researchers. Yarosh and Schueller [2017] and Gobl et al. [2019] both underline that it is important that the researchers learned how to engage and learn from the mental models and views of their co-researchers [66, 123]. Being sensitive to the context and the co-researcher often means that pre-planned methods and approaches have to be adapted on the fly to create a comfortable and stimulating environment for citizens to actively engage in research [61, 78].

## Conclusion

In this scoping review, our main interest was 1) the levels of citizen involvement in current research on technological innovations for health, care or well-being, 2) the used participatory methodologies, and 3) lesson’s learned by the researchers. The increasing number of publications per year in the period 2016-2020, shows the growing interest in citizens science in the field of health, care or well-being. The records that surfaced in this review confirm the diversity of research methods that are applied in citizen science for health care or well-being, mainly applying qualitative methods, often tailored to the specific research problem and group of co-researchers. The five example cases showcase how the contextuality of citizen science as a situated research approach has an impact on the chosen research methods, analysis, and research results. Citizen science in this context requires an open attitude towards how citizens are involved, who the involved citizens are, which methods are used and what kind of topics are researched.

Based on this scoping review, the topic of the required flexibility of researchers in citizen science projects is an opportunity for further research. Although citizen science projects could take more time, it has a meaningful impact on researchers and citizen scientists [Dewa et al. 2020]. Researchers need to engage with co-researchers and find a balance between control and an open structure. The records in this review show some directions to adapt and relinquish control based on learning experience during the citizen science project. Explore this learning process in more depth could provide further understanding and recommendations for researchers as well as citizen scientists involved in citizen science projects. Where is guidance of researchers asked, where is it necessary to give citizens more space, and when would you exert more supervision?

Another follow-up could include exploration of collaboration with vulnerable target groups. The cases and other records in this review show that it is possible to actively involve and collaborate with vulnerable target populations, with or without the use of proxies. Furthermore, based on the minimal findings considering the value of citizen science for the involved citizens, motivation of researchers to start collaborating and motivation of citizens to become active in a project, this could be an interesting direction to explore. Connected to this question on the value of citizen science is the question on the need of a high level of citizen involvement. This scoping review showed that the involvement of citizens in citizen science projects could differ in each research phase. From the included records, it cannot be discussed whether this difference in level of involvement effects the project, research outcome or contribution of the citizens. Despite the lower levels of involvement in one research phase or another, the contribution of citizens in other phases was worthwhile. Therefore, it might be argued that a high level of involvement is not always necessary to execute a valuable citizen science project, but we cannot draw firm conclusions without empirical evidence.

## Data Availability

All data will be available from the TOPFIT Citizenlab database.

## Acknowledgements

The authors thank all colleagues of TOPFIT Citizenlab for their regular support and collaboration. Special thanks to R. Wolkorte and L. Heesink for their involvement at the start of this scoping review and K.E. Konrad and M.E.M. den Ouden for their supervision.

## Author Contributions

**Conceptualization:** Catharina M. van Leersum, Christina Jaschinski, Marloes Bults, Johan van der Zwart

**Data curation:** Johan van der Zwart

**Data analysis:** Catharina M. van Leersum, Christina Jaschinski, Marloes Bults, Johan van der Zwart **Data interpretation:** Catharina M. van Leersum, Christina Jaschinski, Marloes Bults, Johan van der Zwart

**Writing – first draft:** Catharina M. van Leersum

**Writing – review & editing:** Catharina M. van Leersum, Christina Jaschinski, Marloes Bults, Johan van der Zwart

